# Target-dependent parameter-effect landscapes of low-intensity focused ultrasound neuromodulation in humans: a systematic review

**DOI:** 10.64898/2026.09.11.26362849

**Authors:** Hyunwoo Jang, Krishna Swetha Meka, Andrew Yang, George A. Mashour, Anthony G. Hudetz, Zirui Huang

## Abstract

Transcranial low-intensity focused ultrasound (LIFU) offers a rare ability to noninvasively modulate human brain activity in superficial and deep neural structures. Yet the field still cannot reliably predict how a given acoustic protocol will affect a specific brain target. We synthesize 230 LIFU experiments targeting the human cerebrum from 136 articles reporting sufficient acoustic information and neuromodulatory outcomes. We compare associations between acoustic parameters and reported outcomes across cortical and subcortical targets. This reveals divergent parameter-effect landscapes across cortical and subcortical targets. Cortical outcomes follow a nonmonotonic organization across pulse repetition frequency and duty cycle, with distinct zones associated with facilitation, suppression, and null or ambiguous effects. Subcortical outcomes instead vary predominantly along duty cycle, shifting from suppression at lower values toward facilitation at higher values. Intensity and dose factors do not provide a consistent target-independent explanation for outcome polarity, and current evidence is insufficient to identify parameters promoting lasting effects. These findings challenge the assumption that acoustic protocols have generalizable effects across the brain and identify anatomical target as a central dimension that should inform LIFU protocol design. We provide SonoMap, an open, interactive web resource that supports a more systematic, evidence-based approach to human LIFU neuromodulation.

## INTRODUCTION

Noninvasive neuromodulation aims to produce predictable effects in well-defined neural circuits. Transcranial low-intensity focused ultrasound (LIFU) can reach cortical and subcortical structures at sub-centimeter spatial scales^1–4^, and it is less constrained by the depth–focality tradeoff faced by conventional electromagnetic methods^5^. Human LIFU studies have documented both facilitatory and suppressive responses^6,7^. Long-lasting effects have also been reported, with follow-up ranging from minutes to months^6,8,9^. These findings have motivated clinical studies in pain, mood, attention, and consciousness^10–13^.

Currently, LIFU protocol selection remains largely heuristic and guided by precedent, with biological outcomes dependent on multiple protocol parameters. Pulse repetition frequency (PRF) and duty cycle (DC) parameters define how acoustic pulses are distributed in time. Acoustic intensity characterizes the strength of the acoustic field. Dose-related factors such as sonication duration and session count determine how long and how often stimulation is delivered^14,15^. Prior reviews have discussed evidence relating acoustic and dose parameters and neuromodulatory outcomes^16–18^. For example, higher DC has generally been associated with facilitation and lower DC with suppression^4^. However, parameter sweeps in human motor cortex did not show this trend, and outcome polarity also varied with intensity^19,20^. Nominally similar protocols have produced opposite outcomes across studies^21,22^, making the effects of LIFU difficult to predict. These inconsistencies suggest that acoustic parameters alone may not provide a generalizable account of LIFU effects across the human brain.

A central but understudied possibility is that the effect of an acoustic protocol depends on the anatomical target to which it is applied. Across neuromodulation modalities, the same stimulation pattern can produce different effects depending on where it is delivered. For example, direct intracranial electrical stimulation has shown that neuronal responses depend on proximity to white matter^23^. Identical transcranial magnetic stimulation protocols have produced opposite physiological effects across cortical targets^24^. Such target dependence may reflect differences in local cellular composition, circuit architecture, network connectivity, and functional role. Because LIFU can reach both superficial cortical and deep subcortical structures, understanding its target dependence is especially important. A protocol that facilitates activity in one region may suppress activity or produce no measurable effect in another. Thus, determining whether parameter- effect relationships generalize across targets is essential for moving LIFU from precedent-based protocol selection toward a more systematic, precise, and predictive form of human neuromodulation.

Here, we evaluate whether the relationship between acoustic parameters and functional outcomes is shared across, or fundamentally shaped by, human brain targets. We systematically assess 230 human LIFU experiments reported in 136 articles and construct parameter-effect landscapes for cortical and subcortical targets. We first show how outcome polarity is organized across PRF and DC, then determine whether intensity and dose-related metrics provide a consistent target-independent explanation for these outcomes. We also assess how protocol parameters relate to effect persistence. By directly comparing cortical and subcortical evidence, this study evaluates anatomical target as a central dimension of LIFU protocol design. We develop and introduce SonoMap as an open community resource, with an interactive evidence map that helps visualize target- and parameter-dependence of reported human outcomes.

## RESULTS

### Human LIFU literature spans diverse targets with growing subcortical representation

We systematically searched PubMed, Google Scholar, bioRxiv, medRxiv, and Scopus using variants of “low-intensity focused ultrasound” and “transcranial focused ultrasound” (see Methods: Literature search and flow diagram in Fig. S2). The resulting dataset comprised 230 human LIFU experiments (152 cortical and 78 subcortical) from 136 articles published or posted from 2014 through June 2026. Here, an experiment was defined as a distinct target–parameter condition reported within an article. Annual experiment counts increased in recent years (Fig. 1a). The growing number and proportion of subcortical experiments allowed comparison with cortical experiments (Fig. 1b). Parameter selections also shifted over time, with more recent studies using lower PRF and DC (Fig. 1c).

**Fig. 1:**
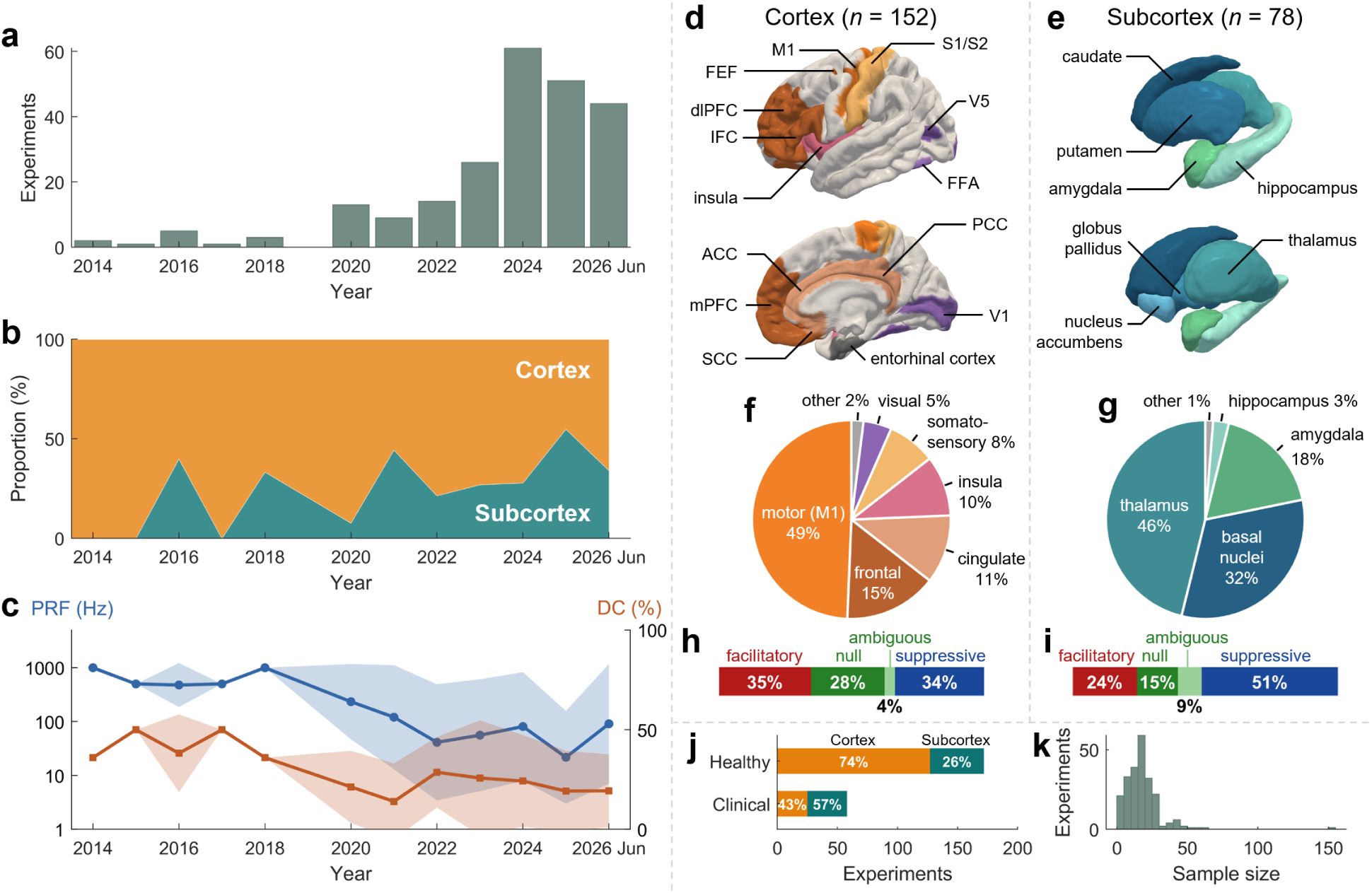
Overview of the human LIFU experiments. (a) Number of experiments reported each year through June 2026. (b) Annual proportions of cortical (orange) and subcortical (teal) experiments. (c) Parameters and annual distributions of PRF (blue) and DC (orange). Lines show yearly means and shaded areas show ±1 SD across experiments. For PRF, means and SDs were calculated after log10 transformation and displayed on a logarithmic y-axis. DC was summarized on the original percentage scale. (d,e) Representative cortical (d) and subcortical (e) target regions included in the dataset. (f,g) Distribution of experiments across cortical (f) and subcortical (g) region categories. See Methods: Region categories for grouping definitions. (h,i) Distribution of facilitatory, suppressive, null, and ambiguous outcomes among cortical (h) and subcortical (i) experiments. (j) Numbers of healthy-volunteer and clinical experiments, partitioned by compartment. Percentages indicate the cortical and subcortical shares within each cohort type. (k) Distribution of reported sample size per experiment. Abbreviations: ACC, anterior cingulate cortex; dlPFC, dorsolateral prefrontal cortex; FEF, frontal eye field; FFA, fusiform face area; IFC, inferior frontal cortex; mPFC, medial prefrontal cortex; M1, primary motor cortex; PCC, posterior cingulate cortex; SCC, subgenual cingulate cortex; S1, primary somatosensory cortex; S2, secondary somatosensory cortex; V1, primary visual cortex; V5, middle temporal visual area. Source data are provided as a Source Data file.

Within each anatomical compartment (i.e., cortex or subcortex), experiments were concentrated in a few target regions (Fig. 1d–g). Primary motor cortex accounted for 49% of cortical experiments, followed by frontal, cingulate, insular, and somatosensory regions. Thalamic targets accounted for 46.2% of subcortical experiments, followed by the basal nuclei and the amygdala.

Outcome polarity distributions differed between cortical and subcortical experiments (*p* = 0.0328, two-sided Pearson’s chi-squared test; Fig. 1h,i). Cortical experiments showed a relatively balanced distribution, with facilitation and suppression occurring at comparable rates. Subcortical experiments, by contrast, were skewed toward suppression. Here, facilitation and suppression denote the inferred direction of change in the physiological function represented by the neural readouts. When neural readouts were unavailable, we used the directional interpretation of the behavioral or clinical outcomes reported by the original authors. These labels describe system-level outcomes and do not imply cellular excitation or inhibition (see Methods: Outcome polarity classification).

Most experiments enrolled healthy volunteers (75%), with a smaller clinical subset (Fig. 1j). Sample sizes were modest (17.3 ± 13.6 participants per experiment, mean ± SD; Fig. 1k) and did not differ across outcome polarity categories (*p* = 0.0685, two-sided Kruskal-Wallis test).

### Cortex and subcortex show distinct response landscapes across temporal parameters

We mapped each experiment in PRF-DC space (Fig. 2) for cortex and subcortex separately. We used Gaussian kernel density estimation to compare the smoothed local densities of facilitatory, suppressive, and null-or-ambiguous outcomes. Regions with total kernel density below 3 were hatched to indicate sparse sampling. Unexplored regions were left blank (see Methods: Parameter-effect landscape generation).

**Fig. 2:**
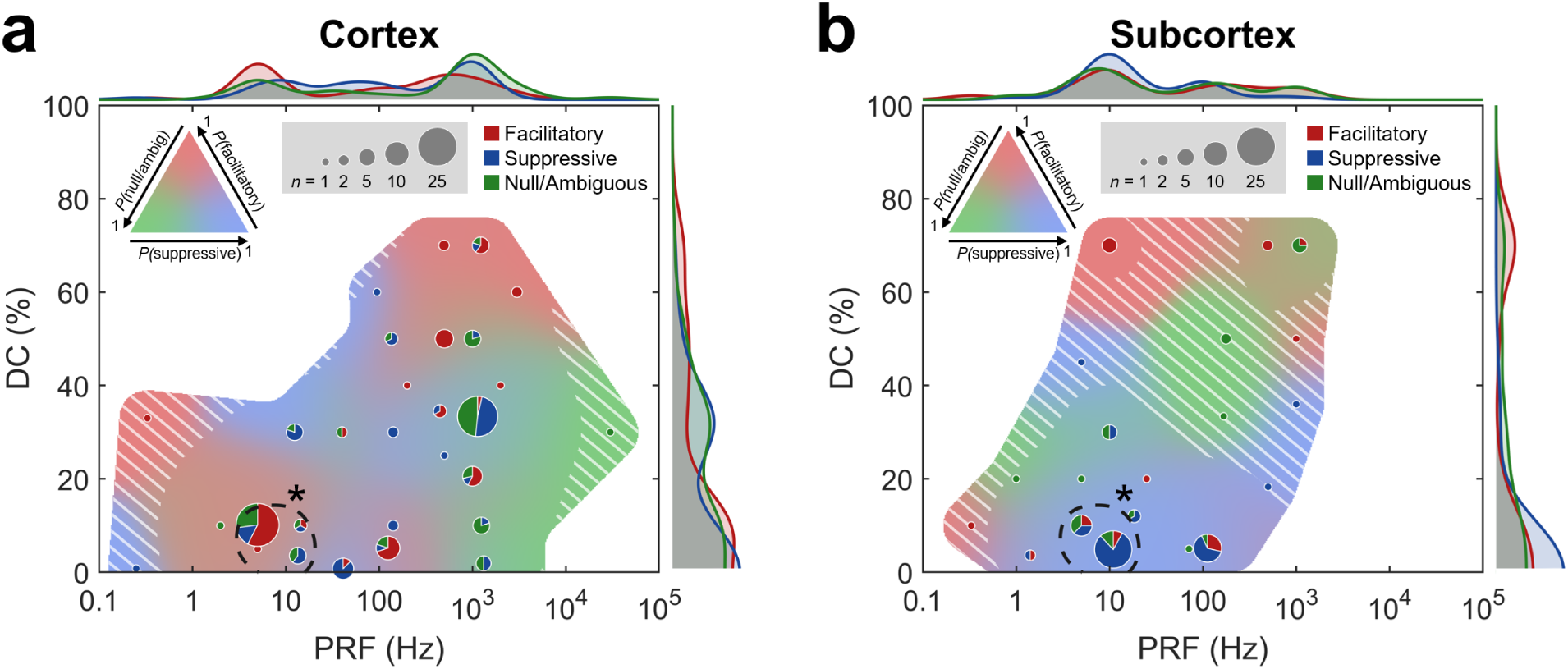
Parameter-effect landscapes in PRF-DC space for cortical and subcortical experiments. (a) The cortical landscape (*n* = 150) shows four broad response zones comprising low-PRF/low-DC facilitation, intermediate-PRF/intermediate-DC suppression, high-PRF/high-DC facilitation, and high-PRF/low-DC null-or- ambiguous outcomes. Two continuous-wave experiments with a DC of 100% were excluded because PRF was undefined. (b) The subcortical landscape (*n* = 78) shows a DC-aligned shift from suppression at lower DC toward facilitation at higher DC. Pie charts show outcome composition within PRF-DC grid bins, with area proportional to the number of experiments in each bin. Red indicates facilitatory outcomes, blue suppressive outcomes, and green null-or-ambiguous outcomes. Background colors show locally smoothed outcome composition estimated by Gaussian kernel density estimation and mapped using the ternary color scale shown in each panel. The smoothed color fields are descriptive and do not statistically define zone boundaries. White hatching identifies sparsely sampled regions with a total kernel density below 3. Dashed outlines and asterisks mark the low-PRF/low-DC zone containing coordinates where outcome distributions differed between compartments (*p*_FDR_ < 0.05, two-sided Freeman–Halton exact tests). Marginal curves show smoothed polarity-specific distributions of PRF and DC. Source data are provided as a Source Data file.

The cortical landscape suggested four broad response zones across PRF and DC (Fig. 2a). Facilitation was most common at low PRF (< 10 Hz) and low DC (< 40%). Suppression predominated across intermediate PRF and DC, broadly spanning PRF values of 10 – 1,000 Hz and DC values of 25 – 50%. At high PRF (> 1,000 Hz), null-or-ambiguous outcomes predominated at DC < 50%, whereas facilitation was more common at higher DC. These visually distinct zones suggest a nonmonotonic cortical pattern.

In subcortex, a simpler pattern emerged along DC (Fig. 2b). At DC < 20%, 65% of experiments were suppressive across a broad PRF range of 1 – 100 Hz, whereas 70% of experiments at DC > 60% were facilitatory. Thus, there was a shift from suppression at lower DC to facilitation at higher DC. But the high-DC boundary remains tentative because sampling at DC > 30% was mostly sparse.

We next tested whether PRF and DC differed across polarity groups within each compartment. For cortex and subcortex separately, we calculated rank-biserial correlations for the three pairwise polarity contrasts in PRF and DC. To account for multiple experiments from the same paper, we obtained 95% confidence intervals using a paper-cluster bootstrap (see Methods: Statistics). All six cortical confidence intervals included zero. Thus, PRF or DC differences between two outcome polarity groups were not robust. In subcortex, suppressive experiments used lower DC values than null-or-ambiguous experiments (*r* = −0.56 [−0.80, −0.26]) and facilitatory experiments (*r* = −0.52 [−0.81, −0.08]). The facilitatory-versus-null/ambiguous DC contrast and all three PRF contrasts included zero. These univariate comparisons support the DC-aligned pattern of the subcortical landscape. Full estimates for the PRF and DC contrasts are reported in Table S1. Sensitivity analyses are included in SI Text: Parameter-effect divergence.

After identifying overall differences, we asked where cortical and subcortical outcomes diverged locally within the PRF-DC space. We tested 58 PRF-DC coordinates that had both cortical and subcortical experiments within the prespecified local neighborhood (see Methods: Parameter-effect divergence). Among these, six showed significant cortical-subcortical differences in outcome distributions (*p*_FDR_ < 0.05, two-sided Freeman–Halton exact test). They were clustered in PRF = 5 – 11 Hz and DC ≤ 10% (dashed outlines in Fig. 2; exact coordinates in Table S2). This overlaps with the canonical theta-burst transcranial ultrasound stimulation (tbTUS) protocol of PRF = 5 Hz and DC = 10%^6,8,25^. Within this zone, cortical experiments more often reported facilitatory outcomes^6,8,25^, whereas subcortical experiments more often reported suppressive outcomes^3,9,26^.

### No intensity or dose-related metric shows a consistent association with outcome polarity across cortex and subcortex

We next asked whether intensity and dose-related metrics were associated with outcome polarity and whether those associations, if any, were consistent across cortex and subcortex. A simple threshold-like relationship would predict null-or-ambiguous outcomes at lower intensity or dose and directional (i.e., facilitatory or suppressive) outcomes at higher values. The primary analysis in Fig. 3 included two intensity metrics and two dose-related factors. The intensity metrics are transcranial spatial-peak pulse-average intensity (*I*_SPPA.tc_; intensity averaged over the active portion of each pulse) and transcranial spatial-peak temporal-average intensity (*I*_SPTA.tc_ = *I*_SPPA.tc_ × DC/100; intensity averaged over the full pulse period). Total sonication time and energy exposure (total sonication time × *I*_SPTA.tc_) were selected as dose-related metrics.

**Fig. 3:**
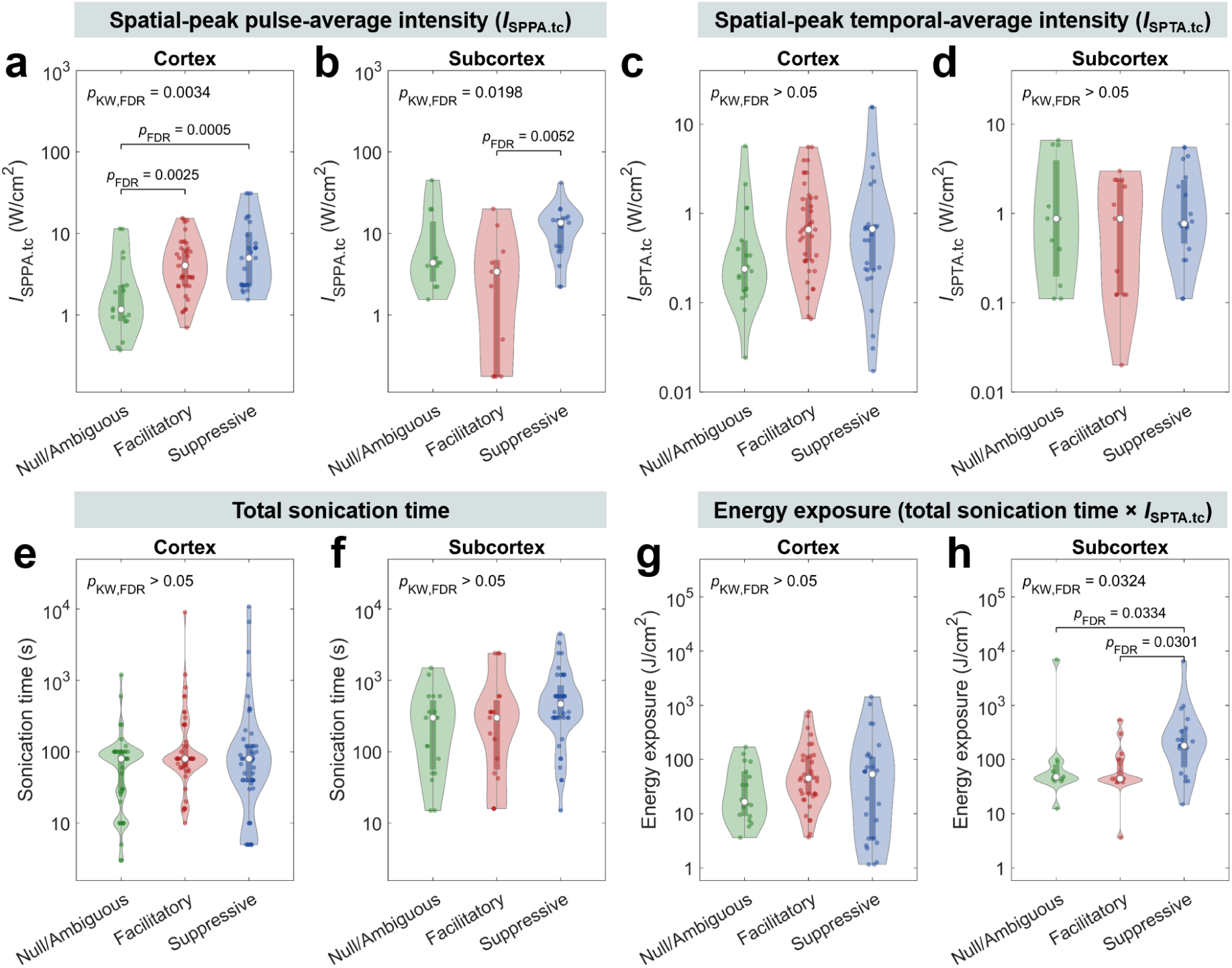
Intensity and dose-related metrics across outcome polarity. Panels are shown separately for (a,c,e,g) cortex and (b,d,f,h) subcortex. Panels show (a,b) transcranial spatial-peak pulse-average intensity (*I*_SPPA.tc_), (c,d) transcranial spatial-peak temporal-average intensity (*I*_SPTA.tc_ = *I*_SPPA.tc_ × DC/100), (e,f) total sonication time across all sessions, and (g,h) energy exposure (*I*_SPTA.tc_ × total sonication time). Points represent experiments, violin plots show distributions, and white dots mark medians. *p*_KW,FDR_ denotes the *p*-value from the two-sided Kruskal-Wallis omnibus test FDR-corrected across the eight panels. *P*_FDR_ denotes the FDR-corrected *p*-value from a two-sided pairwise Mann–Whitney test and is shown only for *p*_FDR_ < 0.05. Experiment counts are *n* = 85 for cortex and *n* = 43 for subcortex in panels a–d, g, and h, and *n* = 151 and *n* = 78, respectively, in panels e and f. Source data are provided as a Source Data file.

Among the four parameters, *I*_SPPA.tc_ showed an omnibus association in both compartments (cortex *p*_FDR_ = 0.0034, subcortex *p*_FDR_ = 0.0198; two-sided Kruskal-Wallis test, Fig. 3a,b). The cortical results of *I*_SPPA.tc_ resembled a threshold-like pattern. Null-or-ambiguous outcomes occurred at lower *I*_SPPA.tc_ than facilitatory (*p*_FDR_ = 0.0025; two-sided Mann–Whitney test) and suppressive outcomes (*p*_FDR_ = 0.0005). Facilitatory and suppressive outcomes did not differ (*p*_FDR_ = 0.2623). However, subcortex did not follow this trend. No comparisons involving null-or-ambiguous outcomes reached significance. Instead, suppressive outcomes occurred at higher *I*_SPPA.tc_ than facilitatory outcomes (*p*_FDR_ = 0.0052).

Energy exposure showed an omnibus association with outcome polarity only in subcortex (*p*_FDR_ = 0.0324, two-sided Kruskal-Wallis test, Fig. 3h), and the pairwise pattern was not threshold-like. Suppressive experiments had greater energy exposure than both facilitatory (*p*_FDR_ = 0.0301, two-sided Mann–Whitney test) and null-or-ambiguous experiments (*p*_FDR_ = 0.0334).

*I*_SPTA.tc_ (Fig. 3c,d), total sonication time (Fig. 3e,f), and cortical energy exposure (Fig. 3g) showed no omnibus association with outcome polarity (all *p*_FDR_ > 0.05, two-sided Kruskal-Wallis test). Overall, we found no evidence of a consistent threshold-like relationship between intensity or dose-related metrics and outcome polarity. See Table S3 for full statistics. Sensitivity analyses are included in SI Text: Intensity and dose-related analyses.

### Effect persistence is incompletely reported, and parameter associations remain inconclusive

Identifying protocols that yield lasting effects is central to the clinical translation of LIFU but the current literature provides inconsistent evidence. We used a one-hour timeframe as a pragmatic threshold to assess outcomes. Effects were classified as persistent if they remained present for at least one hour after discontinuation of the stimulus and as transient if return to baseline was reported before one hour. Healthy-volunteer experiments typically characterize protocol-specific perceptual or motor responses, whereas clinical experiments often evaluate therapeutic change across repeated sessions or extended follow-up, such as changes in affect and arousal. We therefore analyzed associations with lasting effect both in the pooled sample and separately within each population type.

Among the 163 facilitatory or suppressive experiments (i.e., excluding null-or-ambiguous), 40% did not report offline assessment (Fig. 4a). 17% ended study follow-up before the one-hour threshold while the effect remained present, leaving durability indeterminate. Among the remaining 71 experiments, 62% were classified as persistent and 38% as transient (Fig. 4b). Persistence was more frequent in clinical than in healthy-volunteer experiments (75.8% versus 50.0%, odds ratio = 3.12, *p* = 0.0303, two-sided Fisher’s exact test).

**Fig. 4:**
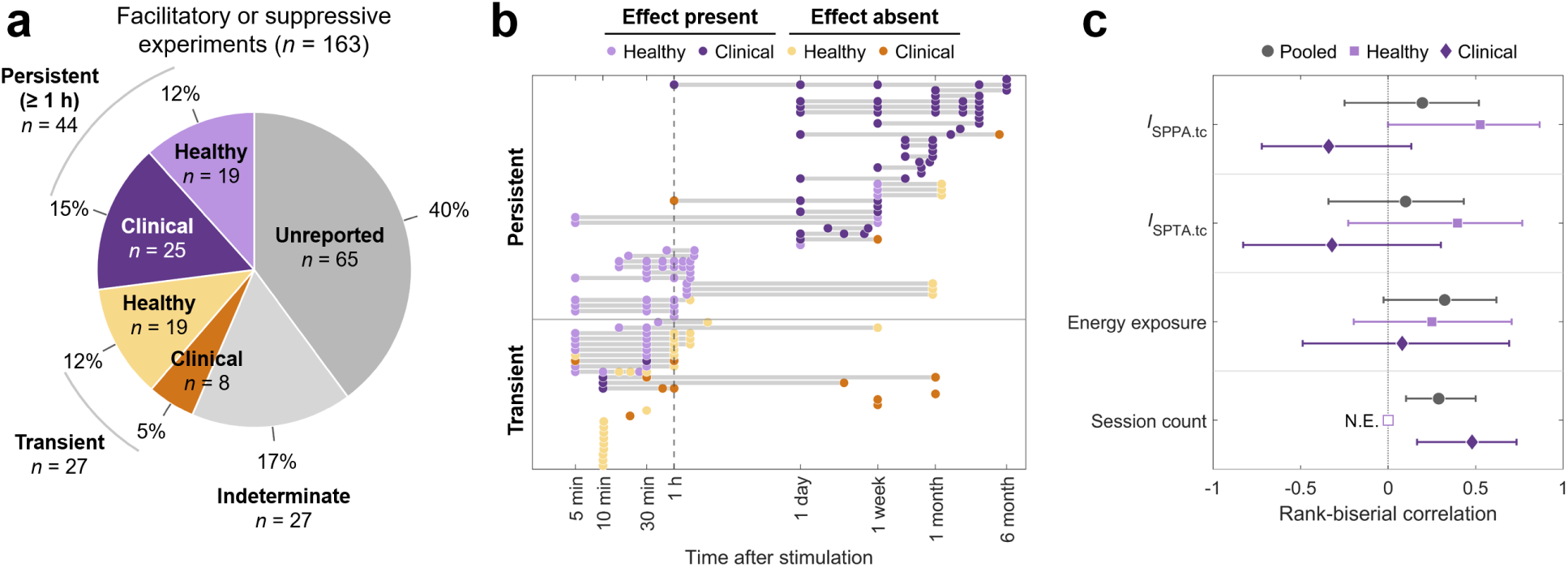
Effect persistence reported in human LIFU literature. (a) Classification of 163 facilitatory or suppressive experiments, with persistent and transient experiments stratified by population type. Using a tentative one-hour threshold, effects were classified as persistent if they were present at or beyond one hour, transient if a reported offline measurement showed absence and no effect was documented at or beyond one hour, indeterminate if reported follow-up ended earlier with the effect present, and unreported if no offline measurement was reported after the final LIFU session. (b) Offline time courses for the 71 experiments with classifiable effect persistence. Each row represents one experiment, and circles mark reported offline measurements. Violet and orange indicate effects present and absent, respectively. Light and dark shades indicate healthy-volunteer and clinical experiments, respectively. The vertical dashed line marks one hour. Horizontal lines of each row connect the first and last plotted offline measurements within each experiment. Rows are ordered by the last time at which an effect was present and then by the last plotted measurement. (c) Rank-biserial correlations for the four displayed parameters. Gray circles, light-purple squares, and dark-purple diamonds show pooled, healthy-only, and clinical-only estimates, respectively. Horizontal lines show 95% paper-cluster bootstrap confidence intervals. Intervals excluding zero indicate associations that were robust to paper-level resampling. Session count was not estimable in healthy experiments because all used one session. *I*_SPPA.tc_ and *I*_SPTA.tc_ denote transcranial spatial-peak pulse-average and temporal-average intensity, respectively. Source data are provided as a Source Data file.

We next compared ten parameters between persistent and transient experiments. These included PRF, DC, *I*_SPPA_ and *I*_SPTA_ in transcranial and free-field forms, sonication duration, sonication time, session count, and energy exposure. None differed after FDR correction in the pooled sample or in either population type (all *p*_FDR_ > 0.05; two-sided Mann–Whitney test). Full statistics are reported in Table S4.

We treated four uncorrected significances as exploratory (Fig. 4c). In the pooled analysis, two parameters were nominally significant. Persistent experiments involved more treatment sessions (*p*_FDR_ = 0.0679, *p*_uncorrected_ = 0.0068; two-sided Mann–Whitney test), and the 95% confidence interval after paper-cluster bootstrapping excluded zero (rank-biserial *r* = 0.29 [0.10, 0.50]). Greater energy exposure was associated with persistence (*p*_FDR_ = 0.1164, *p*_uncorrected_ = 0.0233), but its confidence interval included zero (*r* = 0.32 [−0.03, 0.62]). Regardless of reporting effect persistence, clinical experiments generally had greater energy exposure and more treatment sessions than healthy-volunteer experiments (both *p*_FDR_ < 0.0001; two-sided Mann–Whitney test; Table S5). Thus, the observed associations may partly reflect differences in population type and treatment structure.

Within healthy-volunteer population type, persistent experiments showed nominally higher *I*_SPPA.tc_ (*p*_uncorrected_ = 0.0057; two-sided Mann–Whitney test) and *I*_SPTA.tc_ (*p*_uncorrected_ = 0.0379) but their 95% confidence intervals included zero. In clinical experiments, the corresponding point estimates were negative and nonsignificant.

Overall, no tested parameter showed a robust association with effect persistence. Given the limited number of experiments that assessed offline effects, these analyses remain hypothesis-generating.

### Development of an open, interactive resource to assess parameter-effect landscape

To support target-specific protocol planning, we developed and introduce SonoMap (Systematic Overview of Neuromodulatory Outcomes by Mapping Acoustic Parameters; https://sonomap.github.io), an open- access interactive resource based on the database used in this paper (Fig. 5). SonoMap summarizes the available evidence rather than generating recommended protocols. Investigators can use it to review the precedent protocols and outcomes for a target or parameter range.

**Fig. 5:**
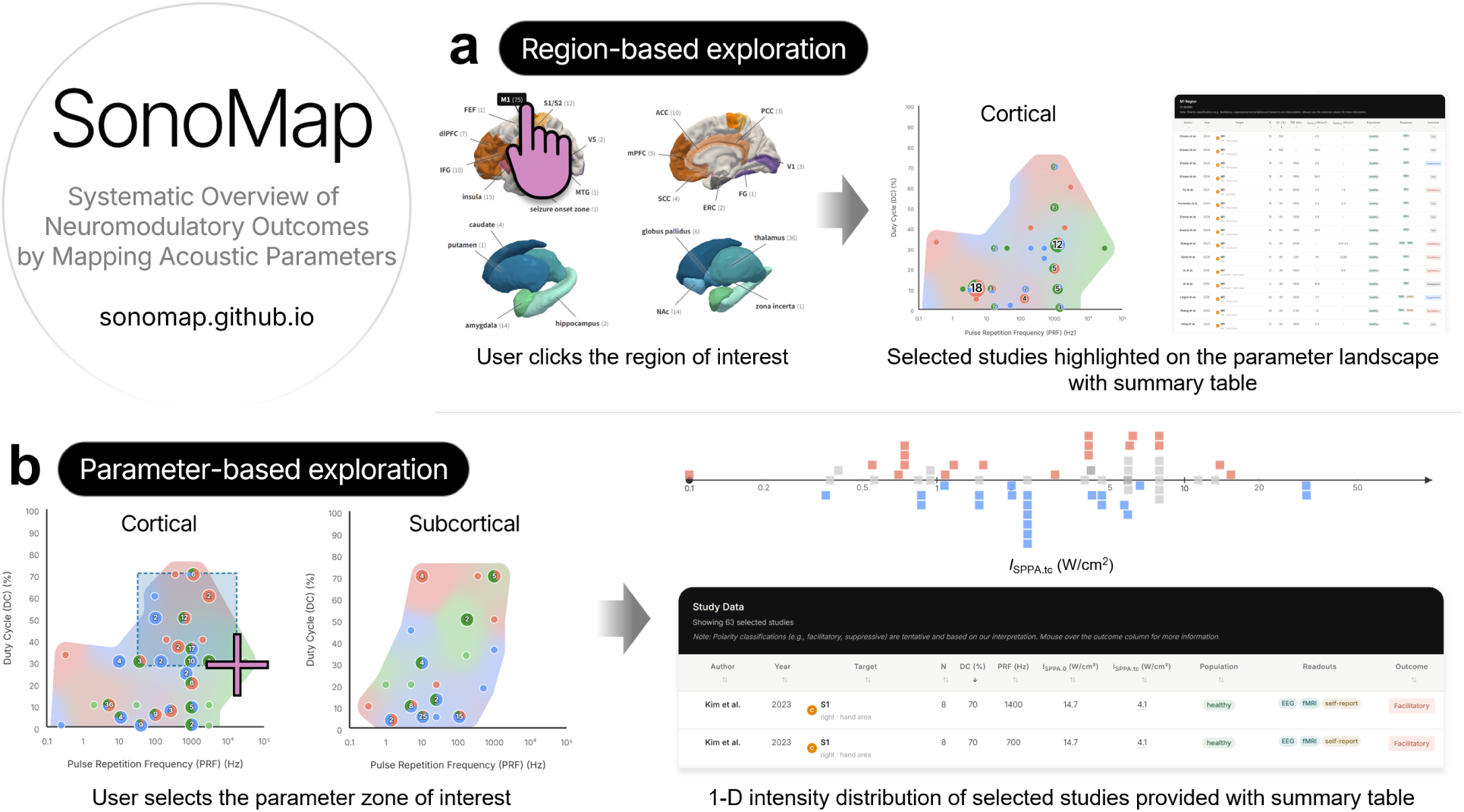
SonoMap, an open and interactive resource for human LIFU literature. The tool’s two main entry points are (a) region-based exploration, in which users select a cortical or subcortical target and view the corresponding experiments on the parameter-effect landscape, and (b) parameter-based exploration, in which users select a region of the cortical or subcortical PRF-DC landscape and view the transcranial spatial-peak pulse-average intensity (*I*_SPPA.tc_) distribution of the corresponding experiments by outcome polarity (facilitatory, red; suppressive, blue; null-or-ambiguous, gray). Both views return a summary table of acoustic parameters, population type, readouts, and detailed outcomes. A separate full-data browser provides direct access to all records. The underlying database is openly accessible, community-contributable, and released under a CC BY 4.0 license (https://sonomap.github.io; code under MIT).

SonoMap offers two main ways to explore the database. The region-based view summarizes all experiments reported for a selected cortical or subcortical target and shows them on the corresponding PRF-DC landscape (Fig. 5a). The parameter-based view retrieves cortical or subcortical experiments from a selected PRF-DC zone. It then plots the outcome polarity of the experiments across the intensity axis (Fig. 5b). Both views are accompanied by a summary table listing sample size, readouts, acoustic parameters, and detailed outcomes. Separating the cortical and subcortical landscapes preserves the compartment-specific divergence reported in this paper. SonoMap accepts community submissions of new human LIFU studies through a dedicated submission page.

## DISCUSSION

Evidence that neural responses to LIFU depend on combinations of acoustic parameters is not new. Animal studies have shown that responses vary with PRF^27^, DC^28^, and intensity^29,30^. Parameter optimization has also identified distinct parameter combinations for excitation and inhibition^31^. However, whether these parameter–response relationships generalize to humans and, if so, whether they are consistent across human brain targets has not been established.

In our synthesis of 230 human experiments from 136 articles, acoustic parameters alone did not account for reported LIFU outcomes across targets (Table 1). First, associations of PRF and DC with outcome polarity differed between cortical and subcortical targets. Second, higher intensity or dose also did not consistently predict outcome polarity across targets. These discrepancies show that existing framework is insufficient to predict outcome polarity at a given human target, and anatomical target should operate as a primary variable in protocol design.

**Table 1:** Summary of main findings and their implications.

| Evidence domain | Main observation | Target-dependence | Implication |
| --- | --- | --- | --- |
| PRF-DC landscape | The global organizations differed between cortex and subcortex. | Cortical outcomes suggested a nonmonotonic organization.<br>Subcortical outcomes showed a stronger alignment with DC. | One parameter rule does not apply to both compartments. |
| Low-PRF/low-DC zone | PRF 5-11 Hz and DC ≤ 10% showed significantly different outcome polarity distributions. | Cortical experiments were largely facilitatory.<br>Subcortical experiments were largely suppressive. | Canonical theta-burst protocol should not be assumed to have the same effect across targets. |
| Intensity and dose metrics | Outcome polarity was associated with pulse-average intensity in both compartments and by energy exposure only in subcortex. | Pulse-average intensity pairwise contrasts differed by compartments. | Current evidence does not support a common intensity threshold or dose rule across targets. |
| Effect persistence | Effect persistence was under-reported.<br>No tested parameter robustly differentiated transient and persistent outcomes. | Target dependence remains unresolved. | No acoustic predictor of persistence is established. Prospective target-stratified follow-up is needed. |

Biophysical models have predicted neural population responses from acoustic parameters based on neuronal intramembrane cavitation^32–34^. In their implementation, inhibitory and excitatory neurons differ in their sensitivity to acoustic pulses. DC and intensity primarily determine the response, whereas PRF has less influence. In general, increasing DC at a fixed intensity shifts the predicted response from net suppression to net activation. Increasing intensity at a fixed DC shifts from no spiking to net suppression and then net activation.

These predictions only partly matched our findings. The DC-aligned trend of subcortex (Fig. 2b) qualitatively resembled the model prediction, but the nonmonotonic cortical PRF-DC pattern did not (Fig. 2a). Intensity also did not reproduce the predicted sequence of null response, suppression, and activation (Fig. 3a–d). This mismatch may arise because the current models only represent a simplified local circuitry, while omitting target-specific circuit architecture and large-scale network embedding.

Cortical microcircuits contain dense recurrent connections between excitatory and inhibitory neurons^35^. In this configuration, the activity of directly stimulated neuronal populations can be altered by local recurrent feedback^36–38^. Population-level outcomes may therefore deviate from the model predictions. Conversely, if local recurrent coupling is weaker, population-level responses may more closely reflect the cell-specific response. This might appear as the DC-dependent polarity transition as shown in subcortex. However, subcortical regions differ widely in their architecture and connectivity and cannot be treated as a single circuit class^39,40^. Animal studies have also found PRF sensitivity in higher-order thalamic nuclei and other deep targets^31,41,42^.

Large-scale network embedding is another candidate source of target dependence. Each brain region occupies different position within the whole-brain network, and participate in distinct systems for gating, arousal, sensory processing, and cognitive control^40,43–47^. For example, posterior cingulate stimulation increased default mode network connectivity, whereas dorsal anterior cingulate stimulation enhanced salience network connectivity in humans^48^. Focal sonication of the lateral geniculate nucleus increased visually-evoked activity in the directly connected ipsilateral primary visual cortex, while ventral posterolateral thalamic stimulation induced more distributed resting-state connectivity increase^49,50^. A local LIFU response may therefore spread through different pathways and networks and produce different functional consequences depending on the target.

In sum, LIFU effects may depend on processes at multiple spatiotemporal scales (Fig. 6a). Acoustic protocol can determine the initial response of certain neuronal populations. Local circuitry and network connectivity may then transform the polarity and detectability of the measured outcome. Target-specific neuroplasticity could add a temporal dimension by determining how the effect decays or stabilizes.

**Fig. 6:**
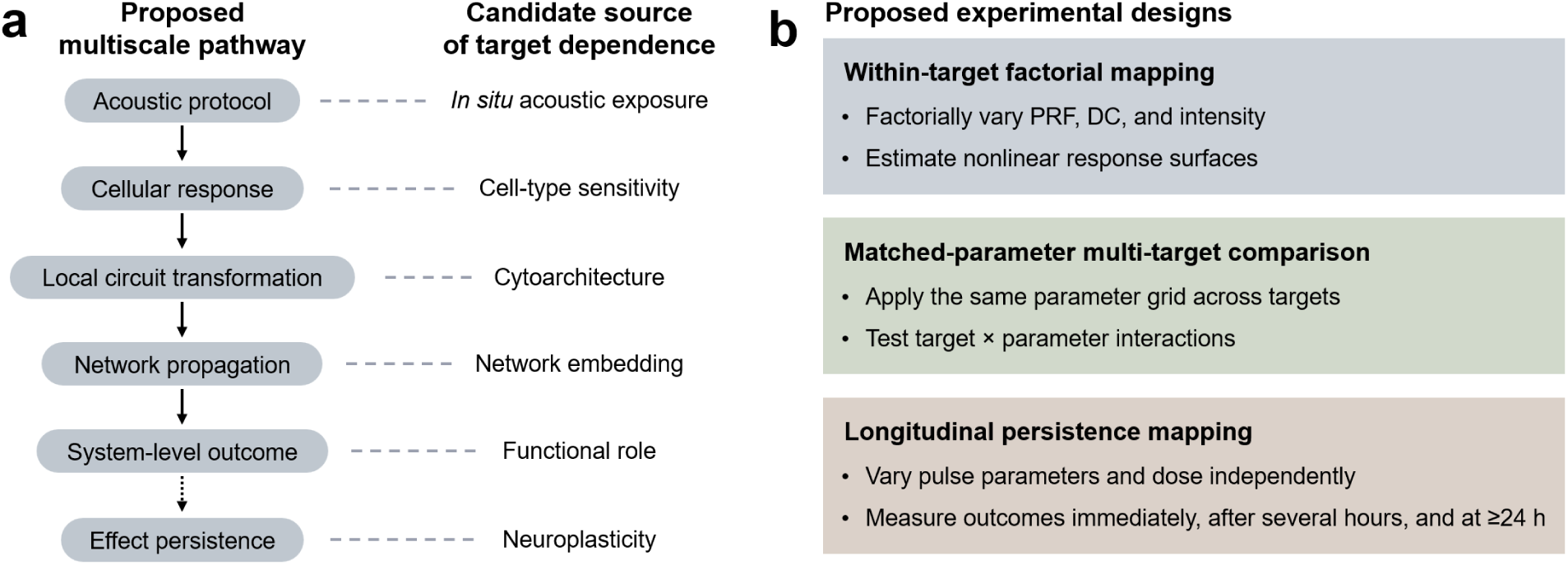
Multiscale framework for target-dependent effects of low-intensity focused ultrasound and proposed experimental tests. (a) Proposed pathway from acoustic protocol to persistent system-level outcome. Solid vertical arrows indicate response transformations across spatial scales. Dotted vertical arrow denotes the temporal scale to effect persistence. Horizontal dashes identify candidate sources of target dependence at each stage. (b) Three experimental structures for testing the framework. Within-target factorial mapping varies pulse repetition frequency (PRF), duty cycle (DC), and intensity to standardize response landscapes. Multi-target comparisons apply a common parameter combination across targets to test target–parameter interactions. Longitudinal mapping identifies predictors of transient or persistent effects by measuring neural outcomes at multiple time points.

This multiscale account can be tested by two complementary experimental strategies (Fig. 6b). First, factorial experiments should map the effects of main parameters including PRF, DC, and intensity within individual brain regions. Our analysis pooled experiments across broad anatomical targets and readouts and a substantial portion of parameter zone remains unexplored. Future studies should vary these parameters across a broad range using a common neural readout. Second, randomized within-subject experiments could apply the same acoustic protocol to various brain targets. The tbTUS protocol (PRF = 5 Hz, DC = 10%) provides a concrete test case because significant cortical-subcortical divergence was already observed in our analysis (dashed outlines in Fig. 2). To distinguish local responses from network- and system-level effects, regional measures should be combined with target-seeded functional connectivity and whole-brain network readouts. These designs would standardize the target-specific parameter–response relationship and allow more accurate comparisons across regions. SonoMap can help prioritize target–parameter combinations where new experiments would most reduce uncertainty.

Both strategies should also assess effect persistence. Although we identified no robust parameter associations, animal studies indicate that ultrasound-induced neuroplasticity depends on pulse-train structure and cumulative exposure, including inter-train interval and total pulse count^51,52^. Future studies should therefore control train structure and cumulative exposure independently. Although we used one hour as a pragmatic threshold, an in vitro study showed that LIFU-induced excitability changes peaked 6 to 8 h after stimulation and diminished by 12 h^53^. Longitudinal protocols should therefore include measurements over the first several hours and again at 24 h or later. This design would distinguish transient from sustained effects and test for parameter-dependent persistence.

This work has limitations. First, outcome polarity is an operational classification and should not be interpreted directly as neuronal excitation or inhibition. The null-or-ambiguous category also combined explicit null effects with mixed or insufficiently resolved outcomes. Second, each experiment is not fully independent, and our cross-study comparisons may reflect shared study context. Experiments from the same group or protocol family can share population types, devices, protocol designs, and study endpoints. Third, binarizing brain regions into cortex and subcortex enabled a first-order comparison but obscured heterogeneity within each anatomical compartment. Fourth, acoustic transmission can contribute to the observed target dependence. Cortical and subcortical targets often differ in skull path, incidence angle, and focal geometry. However, methods for estimating *in situ* acoustic exposure are not yet standardized across studies^54,55^. Finally, null-or-ambiguous outcomes may be under-represented in the literature. Although they were reported more often in recent studies (Fig. S1), and their proportions in healthy-volunteer and clinical experiments did not differ dramatically (32% and 21%, respectively), we cannot exclude publication or reporting bias. Therefore, these results can guide prospective testing but should not be treated as fixed rules for protocol selection.

In conclusion, the available human evidence challenges the assumption of protocol generalizability across brain regions. Acoustic parameters should therefore be selected and interpreted in relation to target anatomy and circuit organization, along with the intended neuromodulatory outcomes.

## METHODS

### Literature search

This review was not prospectively registered, and a formal review protocol was not prepared. Relevant human LIFU studies published or posted through June 2026 were identified through iterative searches of PubMed, Google Scholar, bioRxiv, medRxiv, and Scopus. Each source was last searched on July 2, 2026. Search terms included “low-intensity focused ultrasound human,” “transcranial focused ultrasound human,” “transcranial ultrasound stimulation neuromodulation,” “transcranial focused [ultrasound OR ultrasonic],” “non-invasive ultrasonic stimulation,” “brain ultrasonic neuromodulation,” “low-intensity transcranial ultrasound,” “transcranial [ultrasonic OR ultrasound] stimulation,” “low-energy transcranial focused ultrasound,” “transcranial focused ultrasound stimulation,” “low-intensity transcranial ultrasound,” and “transcranial focused ultrasound,” “transcranial ultrasound TUS.” Eligible preprints were retained to capture recent evidence, while acknowledging that these studies had not yet undergone peer review.

### Eligibility criteria

*Population type.* Healthy volunteers or clinical participants. Animal, in vitro, ex vivo, and purely simulation studies were excluded, as were modeling, hardware, safety verification, or dosimetry papers without human neuromodulation outcomes.

*Intervention.* Non-ablative, low-intensity transcranial focused ultrasound applied with neuromodulatory or therapeutic intent. Studies using high-intensity or ablative focused ultrasound (e.g., lesioning, thalamotomy) without separable low-intensity arms were excluded, as were investigations exclusively targeting non-neuronal effects (e.g., blood-brain barrier opening, ultrasound-mediated drug release). Transcranial pulse stimulation and approaches employing unfocused fields were excluded. Both pulsed and continuous-wave focused-ultrasound protocols were eligible.

*Target.* Only studies applying LIFU to predefined cerebral (cortical or subcortical) gray-matter regions were included. Peripheral, muscle, or spinal applications were excluded, as were studies that explicitly targeted white-matter tracts only. One cerebellar study identified during the search was excluded because the cerebellum falls outside the cerebral cortical-subcortical comparison examined here^56^.

*Acoustic reporting.* Studies lacking sufficient acoustic information to reconstruct intensity and the waveform-relevant temporal parameters were excluded. Pulsed protocols required reporting of both PRF and DC.

Screening of all retrieved articles against these criteria yielded 136 distinct papers meeting inclusion requirements. Fig. S2 shows the PRISMA-based flow diagram of the article selection process^57^. Table S13 enumerates all 136 articles included in the analysis. Table S14 lists examples of excluded articles and the corresponding reasons.

Articles were included only if they met all of the following criteria. K.S.M. and H.J. performed the initial screening against the inclusion and exclusion criteria. H.J. subsequently reviewed both included and excluded articles to verify the eligibility decisions. Reviewers were not blinded to authorship or journal identity.

### Data extraction

Data was manually extracted from each article by either K.S.M. or H.J. All data extracted by K.S.M. were subsequently checked by H.J. against the source. Each eligible article was represented by one or more experiments using a standardized extraction template. Articles with multiple targets or parameter sets were split into multiple experiments with unique target and parameter conditions. Reports including both healthy and clinical participants were additionally split into separate experiments when subgroup-specific outcomes were available. When multiple reported experiments shared identical target and acoustic parameters, they were aggregated into one experiment. The template recorded bibliographic information and participant characteristics, including sample size, demographics, and clinical diagnosis. No sex- or gender-stratified analyses were performed. We extracted the reported target region and hemisphere, PRF where applicable, DC, and free-field and transcranial *I*_SPPA_ and *I*_SPTA_ values where available. When free-field intensity was not reported directly, we derived the corresponding estimates from reported derated intensity or acoustic pressure using the methods described in *Methods: Parameter definitions*.

We also recorded readout modalities, outcomes for each readout, outcome polarity (see *Methods: Outcome polarity classification*), and offline follow-up. Readouts were grouped into neurophysiological and non-neurophysiological measures. Neurophysiological measures included functional magnetic resonance imaging (fMRI), electroencephalography (EEG), magnetoencephalography (MEG), electromyography (EMG)-based motor evoked potential (MEP), functional near-infrared spectroscopy (fNIRS), arterial spin labeling (ASL) perfusion, and magnetic resonance spectroscopy (MRS). Non-neurophysiological measures included sensory, motor, and cognitive task performance as well as self-reported mood, pain, craving, and other clinical symptoms. Outcome polarity was assigned during primary data extraction and subsequently audited against the source text. Uncertain or internally inconsistent cases were manually re-reviewed before the final classification was assigned. Follow-up fields included the reported time points at which the presence of effects was evaluated. We did not formally assess risk of bias and certainty in the body of evidence. The full experiment table, including all extracted parameters, outcomes, and offline follow-ups, are provided in Source Data.

### Classification of cortical and subcortical targets

Each experiment was categorized according to its primary anatomical target. Cortical targets encompassed all regions of the cerebral cortex, including somatosensory cortex (S1, S2), primary motor cortex (M1), visual cortex (V1, V5), lateral frontal areas (dorsolateral and inferior prefrontal cortex), medial prefrontal cortex, cingulate cortex (anterior, anterior midcingulate, posterior, subgenual), entorhinal cortex, and insular cortex. Subcortical targets encompassed deep-brain gray-matter structures, including the thalamus and lateral geniculate nucleus (LGN), basal nuclei (caudate, putamen, nucleus accumbens, globus pallidus, subthalamic nucleus, and zona incerta), amygdala, and hippocampus. Mixed targets involving the ventral capsule together with either the ventral striatum or bed nucleus of the stria terminalis were retained because the designated target included a subcortical gray-matter component. No experiment targeting the ventral capsule alone was included. Classification followed the target explicitly designated by each article. Because LIFU focal volumes are elongated, stimulation could extend into adjacent regions. We did not attempt to model this secondary exposure.

### Outcome polarity classification

To enable cross-study comparison, each experiment was categorized as facilitatory, suppressive, null, or ambiguous based on the physiological or functional interpretation of the primary reported neural, behavioral, or clinical outcome. Given the diversity of experimental readouts, we synthesized outcome polarity rather than pooling effect magnitudes across studies. Classification followed the meaning of the readout rather than its numerical direction alone.

*Facilitatory.* The outcome was interpreted as reflecting greater activation, excitability, responsiveness, or functional output of the relevant system. Representative indicators included increases in motor- or sensory-evoked potential amplitude, increased EEG power, enhanced blood-oxygen-level-dependent (BOLD) signal or functional connectivity, increased perfusion, higher glutamate or glutamine levels, or improvements in task performance.

*Suppressive*. The outcome was interpreted as reflecting reduced activation, excitability, responsiveness, or functional output of the relevant system. Indicators included decreased MEP amplitude, reduced BOLD signal, decreased functional connectivity or EEG power, reduced perfusion, increased gamma-aminobutyric acid (GABA) levels, slower task performance, or decreased sensory experience such as pain.

*Null.* No statistically significant change from baseline or sham was observed.

*Ambiguous.* Polarity was internally inconsistent within a readout modality, or effects were modulatory without clear directionality.

For inferential analyses, null and ambiguous experiments were combined into a single category because neither represented a consistently directional outcome. When polarity differed between neurophysiological and non-neurophysiological readouts, neurophysiological measures were prioritized. Within neurophysiology, central neural signals (fMRI BOLD, MRS) were weighted over peripheral outputs (EMG-based MEP). When direct neurophysiological measures were unavailable, polarity followed the direction and interpretation of the primary behavioral or clinical outcome reported in the source article. Accordingly, these categories do not directly label cellular excitation or inhibition, nor do they encode clinical valence. For example, reductions in pain or craving were classified as suppressive, whereas improvements in coma recovery were classified as facilitatory.

### Parameter definitions

*Pulse repetition frequency* (PRF) is the rate at which ultrasound pulses are delivered, expressed in Hz. *Duty cycle* (DC) is the proportion of ultrasound-on period within each pulse, expressed as a percentage. For rectangular pulses, DC = pulse duration × PRF × 100%. For continuous-wave stimulation at a DC of 100%, PRF was treated as undefined, and these experiments were excluded from PRF-based analyses.

*Spatial-peak pulse-average intensity* (*I*_SPPA_) is the acoustic intensity at the spatial peak averaged over the active portion of each pulse, in W/cm^2^. *Spatial-peak temporal-average intensity* (*I*_SPTA_) is the acoustic intensity at the spatial peak averaged over the full pulse period, in W/cm^2^. It was calculated as *I*_SPTA_ = *I*_SPPA_ × DC/100. The suffixes “.tc”, “.3” and “.0” denote transcranial, derated, and free-field intensity values, respectively. Reported transcranial *I*_SPPA_ estimates served as the primary intensity metric. Free-field values were examined in sensitivity analyses. For studies reporting only conventionally derated intensity (*I*_SPPA.3_), the corresponding free-field intensity was estimated as *I*_SPPA.0_ = *I*_SPPA.3_ × 10^0.3fz/10^, where *f* is the fundamental frequency in MHz and *z* is the estimated propagation depth along the beam trajectory in cm. When only free-field peak acoustic pressure was reported, *I*_SPPA.0_ was estimated assuming a sinusoidal waveform as

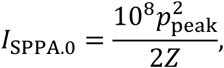

where *p*_peak_ is the pressure amplitude in MPa and *Z* = 1.5 × 10^6^ kg/m^2^·s is the assumed acoustic impedance. The resulting intensity is expressed in W/cm^2^.

*Sonication duration* was defined as the length of one protocol-defined pulse train, including pulse-off intervals. A session was defined as a distinct study occasion during which the planned LIFU protocol was delivered. *Session count* was the number of such occasions per participant. *Total sonication time* was calculated as sonication duration × number of pulse trains per session × session count. It represents cumulative sonication-period time, including pulse-off intervals. *Energy exposure* was calculated as *I*_SPTA.tc_ × total sonication time, expressed in J/cm^2^. It corresponds to time-integrated spatial-peak exposure and does not represent actual energy absorbed by tissue.

### Region categories

For summary visualization (Fig. 1f,g), targets were grouped into broader anatomical categories. Cortical targets were divided into motor (M1), frontal (dorsolateral prefrontal cortex, medial prefrontal cortex, inferior frontal cortex), cingulate (anterior cingulate cortex, anterior midcingulate cortex, posterior cingulate cortex, subgenual cingulate cortex), somatosensory (S1, S2), insula, visual (V1, V5), and other cortical regions (entorhinal cortex, temporal lobe, fusiform gyrus, and mixed-target studies). Subcortical targets were divided into thalamus (including LGN), basal nuclei (caudate, putamen, nucleus accumbens, globus pallidus, subthalamic nucleus, zona incerta, and mixed ventral capsule/ventral striatum targets), amygdala, hippocampus, and other subcortical regions (mixed ventral capsule/bed nucleus of the stria terminalis targets).

### Parameter-effect landscape generation

Parameter-effect landscapes (Fig. 2) were generated per compartment using Gaussian kernel density estimation in log_10_PRF × DC space. Because pulse repetition frequency is undefined for continuous-wave sonication, two cortical experiments delivered at a DC of 100% were omitted from these landscapes. Experiments were placed on a 2D grid spanning log_10_PRF from −1 to 5 (0.1 Hz to 100 kHz) and DC from 0 to 100%. Distances between points were defined using a normalized metric, *d*^2^ = (Δlog_10_PRF)^2^ + (ΔDC/20)^2^, so that *d* = 1 corresponds to a 10-fold difference in PRF or a 20% difference in DC. For each compartment separately, every experiment contributed a standard Gaussian kernel *K*(*d*) = exp(–*d*^2^/2*σ*^2^) to three polarity-specific density surfaces (facilitatory, suppressive, null-or-ambiguous), with bandwidth *σ* set by Silverman’s rule (*σ*_cortex_ = 0.456, *σ*_subcortex_ = 0.437).

At each grid coordinate, the three densities were normalized to local outcome-composition weights. For visualization, each weight was squared and renormalized before weighting a three-color blend (red: facilitatory, blue: suppressive, green: null-or-ambiguous). Experiments were also grouped on a regular grid with 12 bins in log_10_PRF and 10 bins in DC. Both axes used a bin width of 0.5 in scaled space. Each cluster was displayed as a pie chart at its centroid, with area proportional to sample size. To smooth the visual boundary, the explored parameter space was enclosed by a concave hull generated with a shrink factor of 0.6. The hull was expanded by 0.6 units and then contracted by 0.3 units in scaled space. Coordinates outside the resulting boundary were left blank. Within the boundary, coordinates with total kernel density below 3.0 were marked as sparse (white hatching in Fig. 2).

Marginal PRF and DC distributions were estimated using one-dimensional Gaussian kernel density estimation. For each parameter, a single bandwidth estimated from the pooled cortical and subcortical data using the plug-in selector was applied across all compartments and polarity groups (0.242 for log_10_PRF and 7.716 percentage points for DC). DC densities were estimated on bounded support from 0% to 100% using reflection boundary correction.

### Statistics

For each analysis, experiments were selected from the included articles according to the required variables. Parameter-effect landscape analyses excluded continuous-wave experiments because PRF is undefined. Primary intensity analyses were restricted to experiments with a reported transcranial intensity metric. Statistical analyses for effect persistence excluded experiments with null-or-ambiguous outcomes and experiments of which last offline follow-up was within one hour post-stimulation.

Where multiple comparisons were performed, *p*-values were corrected for the false discovery rate (FDR) using the Benjamini–Hochberg procedure. Experiments with missing values for variables required in each analysis were excluded from that analysis, except where transcranial intensity was imputed as described below.

*Descriptive analyses.* The overall difference in outcome polarity distribution between cortical and subcortical experiments was tested with a two-sided Pearson chi-square test. Differences in reported sample size across outcome polarity groups (facilitatory, suppressive, null-or-ambiguous) were tested with a two-sided Kruskal-Wallis test.

*Parameter-effect divergence.* Within each compartment, we quantified differences in PRF and DC among facilitatory, suppressive, and null-or-ambiguous experiments using rank-biserial correlations. For a continuous parameter compared between polarity groups *A* and *B*, the rank-biserial correlation was defined as *r* = 2*P*(*A* > *B*) − 1, with ties counted as 0.5. Values range from −1 to +1, with positive values indicating higher parameter values in group *A*. We calculated three contrasts for PRF and DC: facilitatory-versus-null/ambiguous, suppressive-versus-null/ambiguous, and facilitatory-versus-suppressive. This yielded six correlations per compartment.

To account for multiple experiments from the same paper, we used a paper-cluster bootstrap. Each of 2,000 draws contained the same number of paper clusters as the original compartment-specific dataset. Paper clusters were sampled with replacement, allowing a cluster to be selected more than once or omitted from a given draw. All experiments belonging to a selected cluster were included together and were repeated each time that cluster was selected. Rank-biserial correlations were recalculated in each draw. Point estimates were calculated from the original experiments. Intervals excluding zero were considered robust to paper-level resampling.

Local divergence of the two landscapes at a given PRF-DC coordinate was evaluated using a two-sided Freeman–Halton exact test. Each test included experiments within a radius of *d* = 0.5 in scaled (log_10_PRF, DC/20) space, corresponding to approximately a threefold difference in PRF or a 10% difference in DC. Each 2 × 3 contingency table crossed compartment with the three polarity groups. Coordinates were tested only when the neighborhood contained at least one cortical and one subcortical experiment, yielding 58 testable coordinates among the 73 coordinates represented in the dataset. *P*-values were FDR-corrected across the 58 tests.

*Intensity and dose-related analyses.* Primary analysis of *I*_SPPA.tc_, *I*_SPTA.tc_, and energy exposure was restricted to directly reported transcranial estimates. Within each compartment, metric distributions were compared across the three outcome polarity groups using two-sided Kruskal-Wallis tests. The eight omnibus *p*-values were FDR-corrected. Comparisons that survived this correction were followed by two-sided Mann– Whitney tests of the three pairwise group differences. The three pairwise *p*-values were FDR-corrected within each metric-by-compartment comparison. Paper-cluster bootstrap confidence intervals were used in sensitivity analyses to assess whether the pairwise findings were robust to within-paper dependence.

*Effect persistence.* An offline effect was defined as an effect observed after completion of the final LIFU session. Persistence classification was restricted to facilitatory and suppressive experiments. One hour was chosen as a tentative binary threshold rather than as a validated timescale of neuroplasticity. Experiments were classified as persistent if an effect was documented at or beyond one hour. They were classified as transient if an offline measurement showed return to baseline before one hour and no effect was observed again at or beyond one hour. Persistence was indeterminate when follow-up ended before one hour and the effect was present at the last measurement. Otherwise, experiments were classified as unreported. Only persistent and transient experiments entered the main analyses. The proportions of persistent and transient experiments were compared between healthy-volunteer and clinical experiments using a two-sided Fisher’s exact test.

The exploratory parameter screen compared persistent and transient experiments across ten acoustic and dose-related parameters including PRF, DC, *I*_SPPA.tc_, *I*_SPPA.0_, *I*_SPTA.tc_, *I*_SPTA.0_, sonication duration, total sonication time, session count, and energy exposure. For *I*_SPPA.tc_ and *I*_SPTA.tc_, we used imputed estimates when directly reported estimates were unavailable. The imputation procedure was as follows. When only free-field intensity was reported, missing transcranial intensity was imputed by multiplying the reported *I*_SPPA.0_ and *I*_SPTA.0_ by the median transcranial-to-free-field ratio calculated separately for each compartment. These ratios were 23.33% for cortex (*n* = 69) and 13.33% for subcortex (*n* = 25). Energy exposure was calculated from this imputed transcranial intensity metrics. Free-field metrics included only reported values.

Parameter differences were tested using two-sided Mann–Whitney tests and summarized using rank-biserial correlations. Positive values indicated higher parameter values among persistent experiments. The screen was performed in the pooled sample and separately within healthy-volunteer and clinical experiments. Session count was not estimable in healthy-volunteer experiments because all used one session. FDR correction was applied separately across the estimable parameters in each screen: ten in the pooled sample, nine in healthy-volunteer experiments, and ten in clinical experiments. Parameters with at least one uncorrected *p* < 0.05 across these 29 comparisons were displayed in Fig. 4c. We obtained 95% percentile confidence intervals using the paper-cluster bootstrap described in Methods: Parameter-effect divergence.

To assess whether the pooled findings could reflect population-type differences, we also compared the same ten parameters between clinical and healthy-volunteer experiments. These analyses included all facilitatory and suppressive experiments, regardless of whether persistence was classifiable. Differences were tested using two-sided Mann–Whitney tests, and the ten *p*-values were FDR-corrected together.

## Software

Statistical analyses were performed in Python 3.14. Figures were generated in MATLAB R2024b.

## DATA AND CODE AVAILABILITY

Source data, supplementary information, code for analysis replication, and the SonoMap website will be made publicly available upon publication.

## Data Availability

All data produced in the present study will become public upon publication.

## ACKNOWLEDGEMENTS

This work was supported by the National Institute of General Medical Sciences of the National Institutes of Health (grant R01GM103894; PIs: A.G.H. and Z.H.) and the Center for Consciousness Science, Department of Anesthesiology, University of Michigan Medical School, Ann Arbor, Michigan, USA. The content is solely the responsibility of the authors and does not necessarily represent the official views of the National Institutes of Health.

## AUTHOR CONTRIBUTIONS

Conceptualization: H.J. and Z.H.; Methodology: H.J. and Z.H.; Software: H.J. and A.Y.; Validation: H.J. and K.S.M.; Formal analysis: H.J. and K.S.M.; Investigation: H.J. and K.S.M.; Data curation: H.J. and K.S.M.; Writing–original draft: H.J.; Writing–review and editing: H.J., G.A.M., A.G.H., and Z.H.; Visualization: H.J.; Supervision: Z.H.; Funding acquisition: A.G.H. and Z.H.

